# Improved measurement of racial/ethnic disparities in COVID-19 mortality in the United States

**DOI:** 10.1101/2020.05.21.20109116

**Authors:** Joshua R. Goldstein, Serge Atherwood

## Abstract

Different estimation methods produce diverging accounts of racial/ethnic disparities in COVID-19 mortality in the United States. The CDC’s decision to present the racial/ethnic distribution of COVID-19 deaths at the state level alongside re-weighted racial/ethnic population distributions—in effect, a geographic adjustment—makes it seem that Whites have the highest death rates. Age adjustment procedures used by others, including the New York City Department of Health and Mental Hygiene, lead to the opposite conclusion that Blacks and Hispanics are dying from COVID-19 at higher rates than Whites. In this paper, we use indirect standardization methods to adjust per-capita death rates for both age and geography simultaneously, avoiding the one-sided adjustment procedures currently in use. Using CDC data, we find age-and-place-adjusted COVID-19 death rates are 80% higher for Blacks and more than 50% higher for Hispanics, relative to Whites, on a national level, while there is almost no disparity for Asians. State-specific estimates show wide variation in mortality disparities. Comparison with non-epidemic mortality reveals potential roles for pre-existing health disparities and differential rates of infection and care.

## Introduction

Racial and ethnic disparities in COVID-19 mortality in the United States are attracting intense attention but estimates of their extent differ widely [e.g., 1–2]. With only limited data, the challenge for researchers is to distinguish racial/ethnic direct disparities in infection risk and lethality of COVID-19 from background compositional effects. In this paper, we make new use of demographic methods to address this challenge, providing age-and-place-standardized estimates of racial/ethnic COVID-19 mortality disparities at the national level and for selected states.

Age is recognized as one of the strongest predictors of COVID-19 mortality [3–4]. An accurate assessment of racial/ethnic disparities in per-capita death rates thus needs to account for differences in age structure across groups, as is done for other causes of death [e.g., 5–6]. Percapita rates that do not adjust for age will tend to find Whites with higher mortality rates, as a result of their older age structure relative to non-White groups.

Especially in the early stages of a viral outbreak, there may also be large spatial variation in the prevalence of infection. The early centers of coronavirus infection in the United States have been large cities such as New York, Chicago, Los Angeles, and Seattle—metro regions in which African Americans, Hispanics, and Asians are disproportionately concentrated. Adjustments for geography, the CDC notes, “ensure that the population estimates and percentages of COVID-19 deaths represent comparable geographic areas, in order to provide information about whether certain racial and ethnic subgroups are experiencing a disproportionate burden of COVID-19 mortality” [1].

Place adjustment and age adjustment tend to work in opposite directions and either approach in isolation is potentially misleading. This divergence in results can be seen in Fig. 1 for New York City, the primary center of COVID-19 infection in the United States. Geographic adjustment without age adjustment, as practiced by the Centers for Disease Control and Prevention (CDC) [1, 7],^1^ is shown in panel A. This approach makes disparities seem small among the three largest race/ethnicity groups. The proportion of deaths for Whites (28.2%) is only slightly less than the “weighted distribution of population” (30.5%). The proportion of deaths for Hispanics (also 28.2%) is about the same as the weighted population (28.5%). And Blacks are only slightly overrepresented in mortality (26.8% of deaths versus 23.3% of the weighted population). Overall, it would seem that, compared to Whites, there are no disparities for Hispanics and only a modest disparity for Blacks. On the other hand, age adjustment, as practiced by the New York City Department of Health and Mental Hygiene (DOHMH) and shown in panel B, tells a completely different story. Both Blacks and Hispanics having mortality rates about twice as high as Whites.

**Figure 1.**
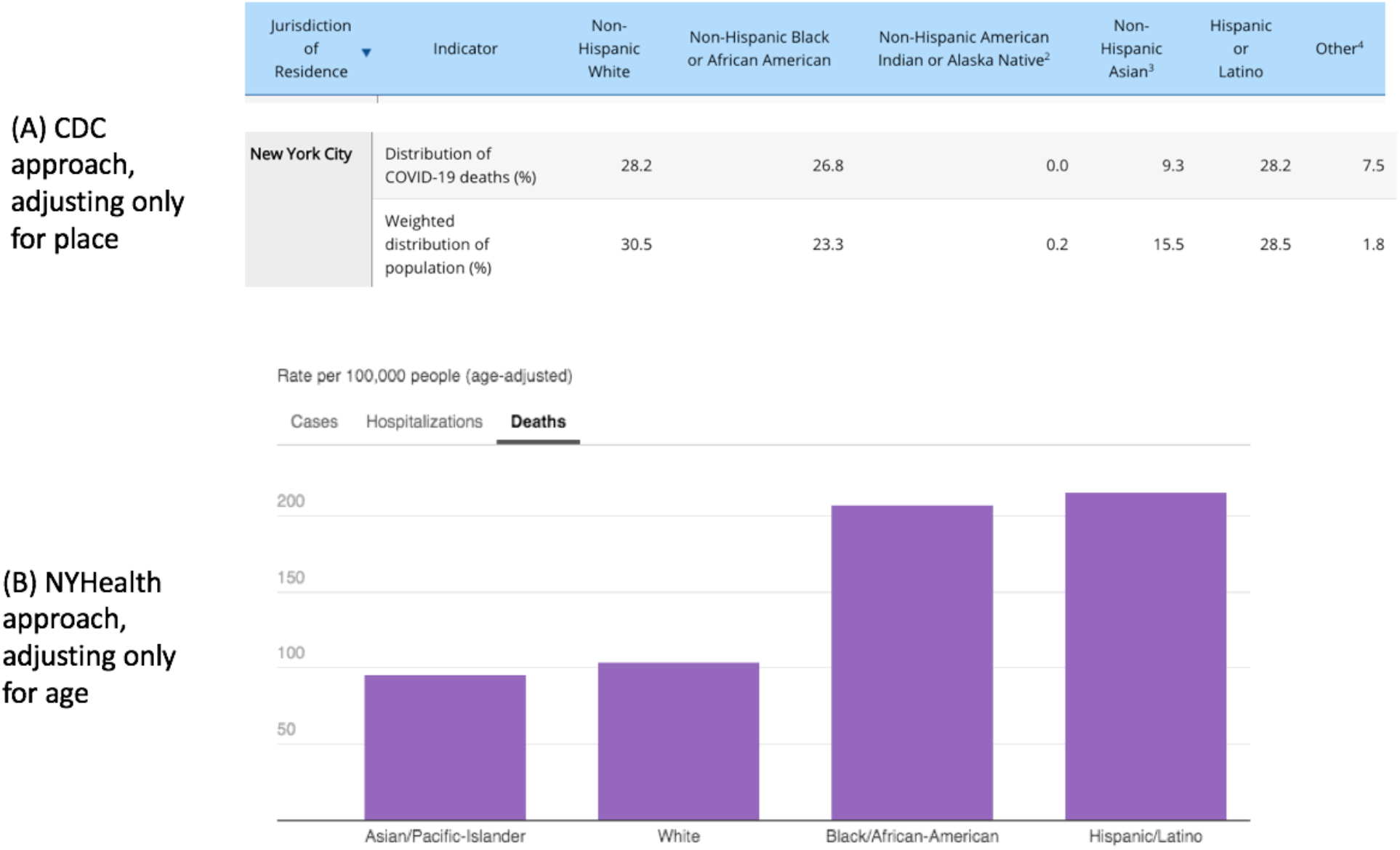
Conflicting indictors of COVID-19 mortality by race/ethnicity in New York City. Mortality adjusted for geographic distribution is represented by panel A, taken from the CDC webpage for provisional COVID-19 death counts by race/ethnicity on May 18, 2020 [1]. Mortality adjusted for age distribution is represented by panel B, taken from the New York City DOHMH COVID-19 webpage [2].

It is possible to resolve such conflicting estimates by adjusting for both place and age at the same time. In this paper, we apply such a method to produce standardized estimates of COVID-19 mortality that are better suited for measuring the magnitude of racial disparities and their causes. Using this approach, we estimate COVID-19 mortality risks for non-Hispanic African Americans (“Blacks”), non-Hispanic Whites (“Whites”), non-Hispanic Asians (“Asians”), and Hispanics of any race (“Hispanics”) nationwide and for selected states.

The procedures we use are designed for limited data and can be implemented widely for estimates at the state and national levels. The absence of detailed death counts that simultaneously report county, age, *and* race/ethnicity of deaths make it impossible to directly compare mortality disparities, but the reporting of the marginal distributions (the number of deaths by age, race, *or* county) allow the use of indirect standardization [8] to adjust per-capita mortality rates for age and geography.

## Methods

The indirect standardization approach is to construct a counterfactual standard schedule of COVID-19 mortality 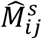 that varies by age *i* and county *j* but is the same for all racial/ethnic groups. A comparison of the observed counts of deaths by race/ethnicity to those predicted by the standard schedule reveals the severity of COVID-19 mortality separately from the effects of age and place. Indirect standardization for both age and place is done by constructing a set of age-specific mortality rates for COVID-19 at the national level. These are then adjusted by level to produce the observed counts of death in each county, to produce the set of standard rates 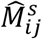. Full details are given in the Appendix; R code for replication is available at https://github.com/josh-goldstein-git/race_covid.

### Data

We use provisional COVID-19 death count data by age, race/ethnicity, and county available online from the CDC [1]. These data summarize the cumulative number of COVID-19 deaths entered into the National Vital Statistics System by the health departments of each state, which in turn compile death records from medical examiners and other authorities at the county level. Because of the coordination required to compile these counts and delays in reporting during the ongoing epidemic, the CDC numbers are explicitly preliminary. In compliance with federal confidentiality requirements, counts are suppressed for counties in which there have been fewer than 10 deaths from the epidemic. As of May 13, CDC reported provisional COVID-19 mortality counts for 322 counties, representing about two-thirds of the national population. CDC’s counts include deaths for which the cause of death is given as COVID-19 as well as deaths for which the coronavirus is identified as the probable cause. Our method does not require counts to be complete, but it does depend on the reliability of the distributions by age, race/ethnicity, and place as given by the CDC.^2^ We use U.S. Census Bureau estimates of population counts by county, race/ethnicity, and age. The most recent detailed population estimates are from 2017.

## Results

Unadjusted per-capita death rates (relative to Whites) are shown at the top of Figure 2 under “Crude Rates.” Taken at face value, they suggest African Americans across the U.S., as a group, have more than 50% greater risk while Hispanics and Asians have about 25% lower risk, relative to Whites. However, as previously discussed, disparities in unadjusted crude death rates are confounded by differences in age structure and the geographically localized nature of the early stages of the epidemic. Accounting for the county-level geography of COVID-19 mortality reduces disparities for Blacks to about half of the level of the per-capita rates (from 56% greater risk to 24% greater risk), as can be seen in the “place standardized” portion of the figure. Place-standardization also reduces the relative death rate for Hispanics slightly and Asians substantially. Age standardized risks are shown in the next section, giving dramatically different results, with Blacks now having a mortality risk of 2.7, relative to Whites, while risks for Hispanics and Asians grow to 1.9 and 1.25, respectively. The disparities increase when the younger age structure of each of these groups, relative to Whites, is considered.

**Figure 2.**
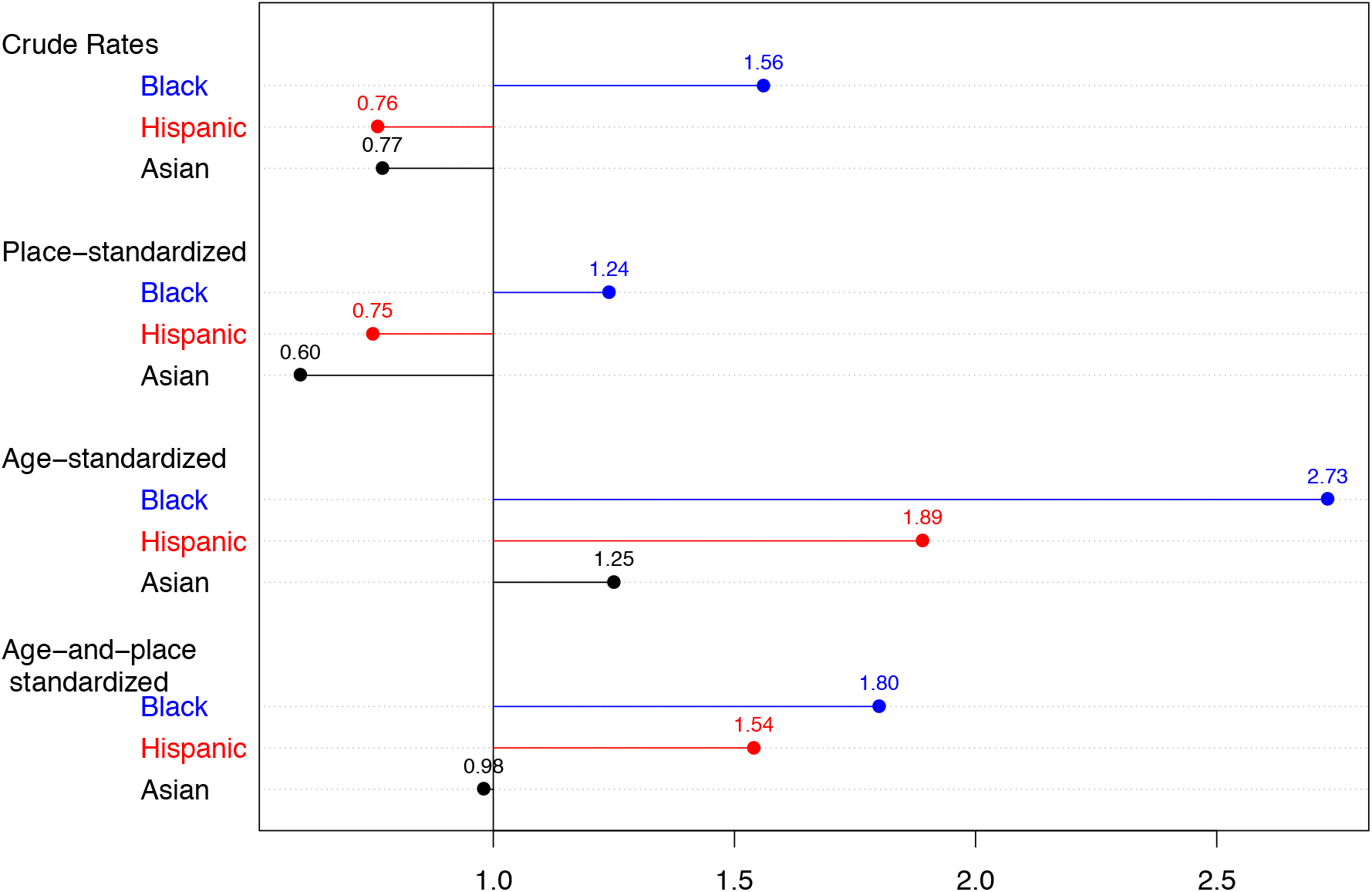
COVID-19 mortality risks by race/ethnicity, relative to Whites, by method. Crude rates are unadjusted per-capita. Place-standardized ratios use indirect standardization to adjust for the geographic composition of each group, by county. Age-standardized ratios use indirect standardization to adjust for the age structure of each group. Age-and-place standardization adjusts for both factors simultaneously. Based on CDC mortality data current to May 13, 2020, including ascertained and probable COVID-19 deaths for counties with 10 or more deaths and 2017 U.S. Census Bureau estimates of county-level demography. Blacks, Asians, and Whites are non-Hispanic, and Hispanics are of any race.

Finally, the bottom of Figure 2 shows age-and-place-standardized relative rates. The COVID-19 mortality risk for African Americans is now 80% greater than for Whites, which is higher than the crude rate and the place-standardized rate, but lower than the age-adjusted rate. For Hispanics, age-and-place standardization turns a place-adjusted Hispanic COVID-19 mortality advantage into a mortality risk 54% higher than for Whites. For Asians, age-and-place standardization nearly eliminates their mortality advantage over Whites, bringing them near parity. Place and age adjustments clearly act in opposite directions at the national level.

Estimated racial/ethnic disparities by state are shown in Figure 3. For African Americans, the South (MS, LA, AL, SC) and Midwest (WI, IL, MI) have high excess mortality, but the Northeast (NY, NJ, MA) has smaller disparities. For Hispanics, the West (NV, AZ, WA, CO, CA) has high excess mortality, while Florida is notably lower.

**Figure 3.**
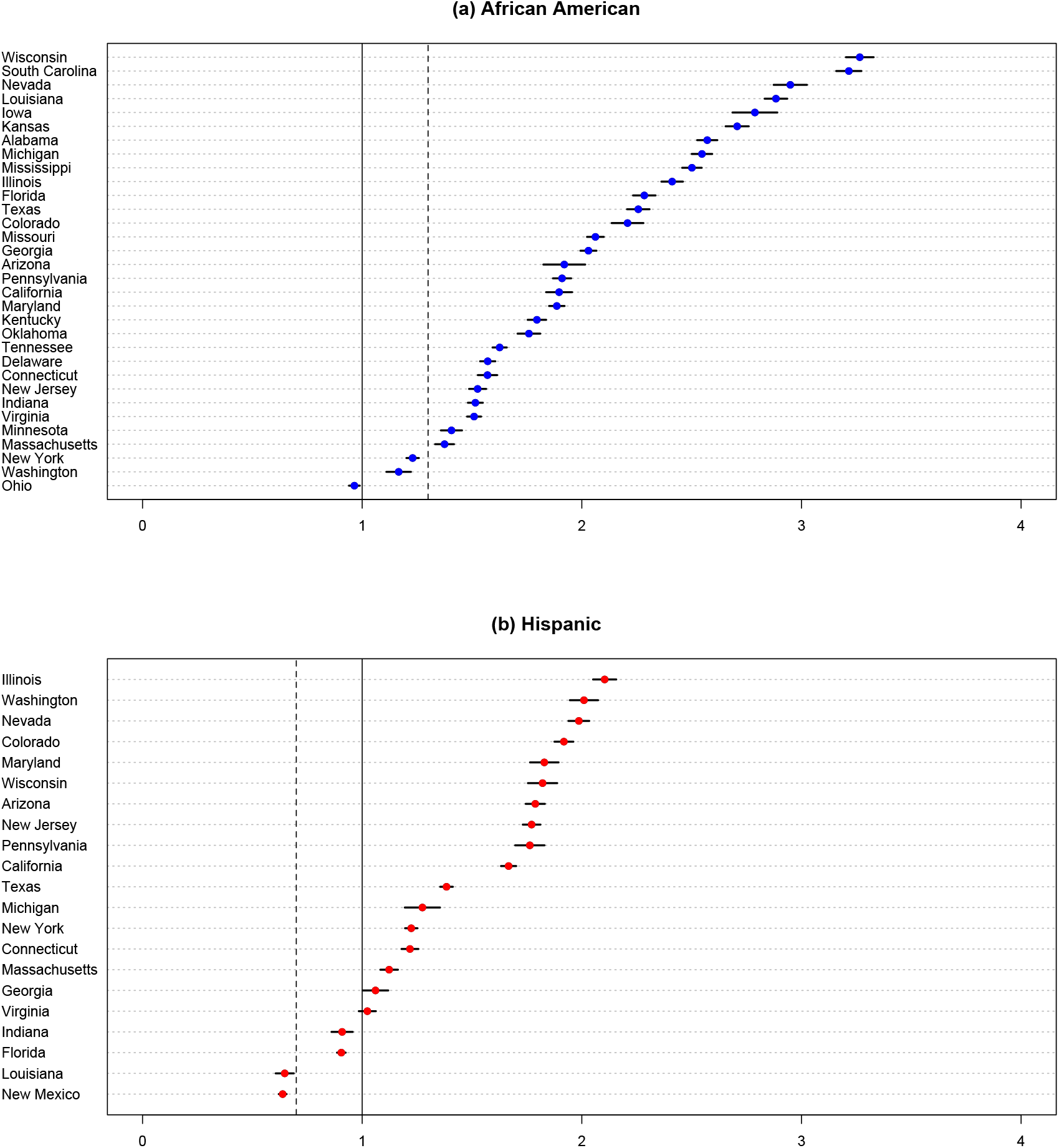
COVID-19 mortality relative to Whites, adjusted for age and place, by state. The solid vertical line at 1.0 is the COVID-19 relative rate for Whites. The dashed vertical line is the approximate national all-cause mortality levels relative to Whites for (a) Blacks/African Americans and (b) Hispanics. States are included with sufficient numbers of cases for estimation. Estimates are based on CDC provisional data updated to May 13, 2020.

## Discussion

We can compare the race/ethnicity risks from coronavirus-specific mortality with the risks from all-cause mortality in non-epidemic times to reveal the role of underlying health conditions inasmuch as these are responsible for previously identified patterns of mortality by race/ethnicity [e.g., 9–11]. U.S. life expectancy estimates imply, relative to Whites, about 30% higher all-cause mortality across all ages for Blacks and about 25% lower all-cause mortality across all ages for Hispanics. This means, for Blacks, that about half of the disparity in COVID-19 mortality (i.e., 0.3 of the 0.7) is potentially attributable to “normal” mortality disparities. For Hispanics, who typically have lower mortality, the coronavirus epidemic appears to have had an even more dramatic effect, completely reversing the so-called “Hispanic paradox” [12–13].

The finding that mortality disparities during the COVID-19 epidemic by race/ethnicity are larger than in non-epidemic times suggests factors other than underlying health disparities are responsible. Principal among these is the differential risk of infection related to exposure at work, in transportation and at home. Differential access to healthcare may also play a role.

Mortality adjustments that take into account the geographic distribution of the epidemic should be considered lower bounds on the true level of inequality. Standardizing for county of residence removes some of the relationship between race/ethnicity and place from analysis, although it leaves a role for within-county differences and allows comparison across states. Standardization is used to remove the effects of “‘extraneous’ influences” [14, p. 24]. Although it is tempting to assume that the magnitude of the COVID-19 epidemic in disproportionately non-White cities like New York and New Orleans may have had more to do with these cities being travel destinations than with their racial/ethnic composition, the high rates of infection within other majority-minority geographies, including Detroit, Milwaukee, and Chicago’s Cook County, seem inextricably linked to the broader history of racial inequality and housing discrimination in the United States [15–16].

As the epidemic spreads, differences due to the initial geographic seeding of the virus in the population may become less important, it may be less advisable to standardize by geography, or to present both the age-adjusted and the age-place-adjusted measures. Finally, state-level variation in disparities may help us to understand the causes of disparities in COVID-19 death rates. We find that the highest excess mortality for Blacks is the Deep South and the Upper Midwest, while disparities are smallest in the Northeast and West. Understanding the reasons for this pattern are an important topic for future research.

This paper offers a method for more refined measurement of racial and ethnic disparities in COVID-19 mortality using limited data. Indirect standardization is a powerful tool that can be widely adopted for the reporting of mortality that adjusts for both age and place. We believe this approach is superior to the one-sided adjustment procedures currently in use by state and local agencies as well as at the federal level at the CDC.

## Data Availability

Publicly available data. Replication code and data available at https://github.com/josh-goldstein-git/race_covid

## APPENDIX Standardization of COVID-19 mortality disparities by race/ethnicity

We estimated Standardized Mortality Ratios (SMR) for each race/ethnicity group (race) by aggregated spatial unit (e.g., a state or the country as a whole). Our estimates take into account, for each small area spatial unit (e.g., counties within a state), the age composition by race and the average severity of the epidemic across all races.

The SMR we calculate is

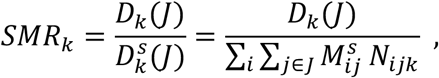

where

*D*_*k*_ (*J*) are the observed counts of COVID-19 deaths of race *k* in aggregated spatial unit *J*,

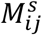 is our standard schedule of COVID-19 per-capita mortality rates by age group *i* and small-area spatial unit *j*, and

*N*_*ijk*_ is the observed census count of the population in age group *i*, in small area spatial unit *j*, of race *k*.

We report the SMRs of each group *k*, relative to that of Whites (*W*), denoting these *R*_*k*_ (*J*), such that

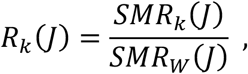

We construct the standard schedule 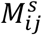 by assuming independence between small area *α*_*j*_ severity and the age schedule of COVID-19 mortality *M*_*i*_.

First, we calculate age-specific per-capita mortality *M*_*i*_ for age group *i* for the United States as a whole,

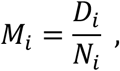

where the *D*_*i*_ are based on provisional COVID-19 death counts from the Centers for Disease Control and Prevention and the *N*_*i*_ on U.S. Census Bureau population estimates.

Then we compute age standardized indices *α*_*j*_ of the intensity of epidemic mortality by small area *j* based on our observed counts *D*_*j*_. Since, by independence, the per-capita rates are the product of age schedule and small area intensity,

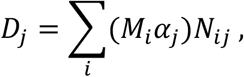

re-arranging gives us an estimate of the age standardized intensity for each small area,

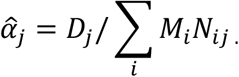

We then, assuming independence, construct our standardized schedule as

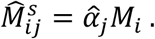

Standardized Mortality Ratios by race give us an estimate of relative mortality, assuming that, within aggregated spatial units, mortality that is specific to age, place, and race is the product of independent terms for age *M*_*i*_, small area spatial unit *α*_*j*_, and race *θ*_*k*_, such that *M*_*ijk*_ = *M*_*i*_*α*_*j*_*θ*_*k*_. Under this assumption, observed deaths of race *k* in aggregated spatial unit *J* is given by

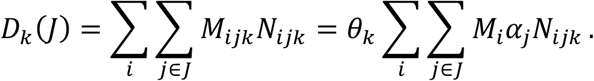

Substituting into (1) shows that *SMR*_*k*_ (*J*) = *θ*_*k*_.

## Footnotes

1 The CDC’s method is to reweight the exposed population to create “weighted population distributions… [to] ensure that the population estimates and percentages of COVID-19 deaths represent comparable geographic areas” [1]. Reweighting has a similar effect to the indirect standardization method we use (see Methods section).

2 Potential reasons that the reported distributions by race, county, and age could vary from the actual distributions include differential timing of reports from hospitals, institutions such as prisons, old age homes, and nursing homes, and at-home deaths. We do not know of research on this issue.

